# A comprehensive descriptive analysis of hip and knee radiographic osteoarthritis in the UK Biobank in relation to joint pain, joint site interrelationships, obesity, race and deprivation: Findings from 59,475 individuals

**DOI:** 10.64898/2026.03.15.26348416

**Authors:** A Hashmi, S Scott, M Jung, FR Saunders, R Ebsim, JS Gregory, L Arbeeva, AE Nelson, NC Harvey, C Lindner, RM Aspden, T Cootes, JH Tobias, BG Faber

## Abstract

**Objectives:** Patients with osteoarthritis (OA) affecting multiple joints have poorer health outcomes than those without, yet most research examines isolated joints, leaving a gap in multi-joint disease. This study aimed to describe radiographically defined hip (rHOA) and knee OA (rKOA) within UK Biobank (UKB), exploring interrelationships across joints, and associations with joint pain, obesity, race and deprivation.

**Methods:** Automated machine learning was applied to left and right hip and knee dual-energy X-ray absorptiometry scans. Radiographic OA (rOA) was defined as custom grades ≥2. Joint pain was assessed through self-reported questionnaires. Descriptive statistics summarised the population characteristics. Logistic regression models examined bilateral and cross-joint associations, as well as associations with joint pain. Adjustments were made for age, sex, race, height, weight and deprivation. Other models examined the associations between body size and OA.

**Results:** Among 59,475 individuals (mean age 65 years; 52.8% female), rHOA prevalence was 4,098 (6.9%)) and 4,841 (8.1%) for the right and left joints, respectively. The corresponding estimates for rKOA were 3,750 (6.3%) and 4,220 (7.1%). Overall, increasing grades of rOA and number of joints affected were more strongly associated with joint pain. Regarding joint-interrelationships, bilateral associations were stronger at the knee, whereas cross-joint associations (hip-knee) were weaker. Associations with BMI and height differed between the hip and knee.

**Conclusions:** Radiographic hip and knee OA exhibit distinct patterns of interrelationship, associations with symptoms and risk factors, suggesting heterogeneity in disease process and the need for joint-specific treatment.

**Key Messages:** *What is already known on this topic?:*

- Osteoarthritis (OA) commonly affects the hip and knee and is associated with pain and disability, with recognised risk factors such as age, obesity and deprivation.
- Increasing interest in multi-joint OA challenges the traditional concept of lower-limb OA as a monoarthritis, but most research examines joints in isolation.
- Genetic evidence suggests that hip and knee OA may differ in underlying mechanisms, yet population-scale comparisons are limited.

*What this study adds?:*

- Among 59,574 individuals, this study identifies that radiographic OA captures structurally and clinically relevant disease with increasing severity and greater number of joints affected, positively associated with chronic joint pain.
- Radiographic hip and knee OA demonstrated strong bilateral but weaker cross-joint associations, indicating preferential within-joint symmetry.
- Risk factors differed by anatomical site with BMI and weight strongly associated with knee OA and weakly associated with hip OA. Height showed the opposite associations.

*How this study might affect research, practice or policy?:*

- These findings support that hip and knee OA may partially represent different disease processes rather than a single condition.
- Clinical practice should consider cumulative joint involvement and joint-specific risk factors.
- Future research should consider the development of more targeted treatment to prevent multi-joint progression.

## Introduction

Osteoarthritis (OA) is a painful, chronic condition that limits mobility and activities of daily living, contributing substantially to disability worldwide(^1^). It affects over 565 million people globally, with prevalence projected to rise by 60-100% by 2050, due to an aging population and rising rates of obesity, the latter of which is estimated to account for approximately 20% of OA cases(^2^). Beyond age and obesity, OA has a diverse risk factor profile with multiple contributors influencing its progression, including genetics, joint injury, deprivation and race. For example, rates are higher in more deprived areas(^3^) and Black-Americans experience a greater burden of knee OA, with higher rates of both radiographic changes and symptoms(^4^). However, given the aforementioned epidemiological shifts, there remains a need for up-to-date estimates derived from contemporary population cohorts.

Historically, lower-limb OA has largely been considered an asymmetric monoarthritic disease, but recent research has introduced the concept of multi-joint OA, where more than one joint is affected. It has been shown that those with OA affecting more than one lower limb joint have poorer health outcomes and increased pain(^5^). Complementary research has challenged traditional assumptions that disease progression is driven solely by degeneration of the articular joint surface, proposing instead that OA should be considered a systemic disease, which fits with the concept of multi-joint OA(^6^). Studying OA across multiple joints within the same individual is therefore vital in understanding the whole-body nature of disease patterns and progression.

Furthermore, new insights from genetic studies have suggested that OA is not a single homogenous condition(^7^). The largest genome-wide association study shows that genetic loci confer risk differentially across joint sites, indicating that OA at different anatomical locations may have distinct underlying biological mechanisms(^8^). If this were the case, then systemic risk factors such as obesity might influence osteoarthritis risk differently across joint sites. In addition, the extent to which OA in one joint influences the development of OA in another site might vary.

UK Biobank offers a unique opportunity to investigate prevalence and predictors of multi-joint OA at scale, as hip and knee high-resolution dual-energy X-ray absorptiometry (DXA) scans, which are akin to routine radiographs, are available(^9^). Previous studies have demonstrated that radiographic features of OA can be derived from DXA scans and that these measures are clinically relevant(^10, 11^). The aim of this study was leveraging data from UK Biobank, to i) undertake a descriptive analysis of radiographically defined hip and knee OA, ii) confirm their associations with joint pain, iii) examine the interrelationship of OA across the hip and knee joints, iv) explore associations with body mass index (BMI), height and weight, and v) identify any differences according to deprivation and race.

## Materials and Methods

### Population

UK Biobank is a longitudinal cohort study that aimed to recruit a population of ∼500,000 individuals aged 40-69 between 2006-2010, from across the UK. Participants underwent extensive genetic and physical phenotyping and completed questionnaires on socio-demographic, lifestyle and environmental factors(^12^). Initiated in 2014, the extended imaging study sought to gather medical imaging data, including hip and knee DXA scans, from 100,000 UK Biobank participants(^13^). UK Biobank obtained ethical approval from the National Information Governance Board for Health and Social Care and Northwest Multi-centre Research Ethics Committee (11/NW/0382). All participants provided informed consent for the collection and use of their data and for follow-up via linkage to electronic healthcare records.

### DXA-derived radiographic osteoarthritis

#### Image Analysis

High-resolution bilateral hip and knee DXA scans were obtained using Lunar iDXA scanners (GE Healthcare, Madison, WI, USA), with participants laying supine. Validated machine-learning algorithms, previously trained on large manually annotated datasets, were applied to these scans to delineate joint shape and to characterise the features of rOA (Figure 1)(^10, 11^). Measurements included osteophyte area and joint space width, while subchondral sclerosis and cysts were not included due to their infrequency on DXA imaging. All images underwent manual review to verify the automated measurements and osteophyte segmentation.

**Figure 1.**
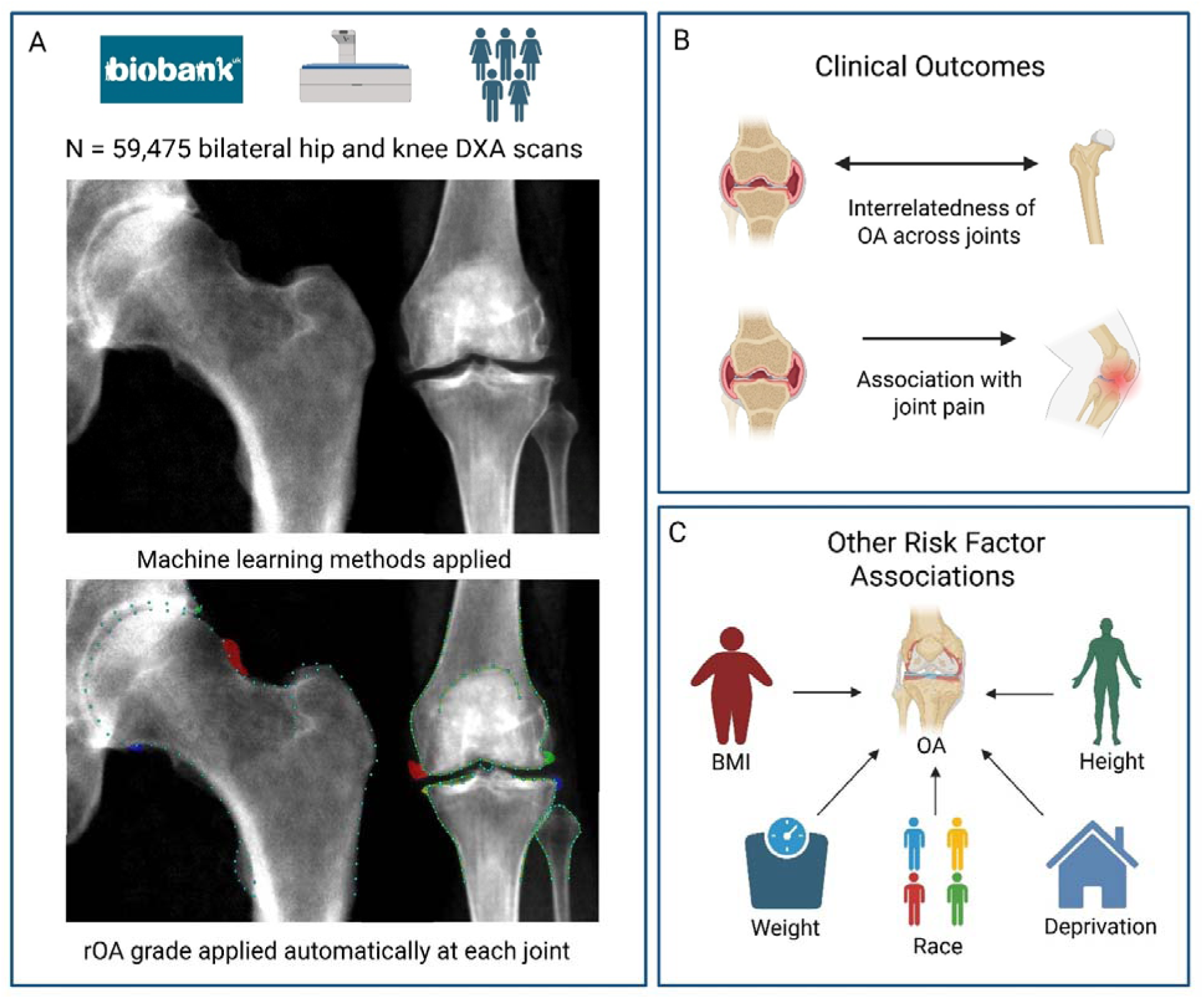
Visual summary of the methodology used in this study A: Machine learning methods were applied to bilateral hip and knee DXA scans from 59,475 UK Biobank participants. Point-placement was used to deri minimum joint space width (mJSW). Osteophytes were identified through shaded colours. Semi-quantitative grades for osteophytes and joint space narrow were derived from osteophyte area and mJSW and combined to form radiographic osteoarthritis grades. B: Interrelatedness of OA across joints and associations with joint pain. C: Associations with BMI, height, weight, deprivation and racial background.

#### Generation of radiographic osteoarthritis

Osteophytes were semi-quantitatively graded 0-3 for each anatomical joint based on their area (mm^2^). The hip osteophytes were assigned at the lateral acetabulum, the superolateral femoral head and the inferomedial femoral head(^14^). Knee osteophytes were graded at the medial and lateral femur and the medial and lateral tibia. Minimum joint-space width measurements were obtained at the superior hip and medial knee compartments. These were used to grade joint-space narrowing from 0-3 as previously published(^11, 15^). An overall radiographic OA (rOA) score was created for each joint by summing grade scores for osteophytes and joint-space narrowing. Grades of radiographic hip OA (rHOA) were assigned using thresholds: grade 0 (sum = 0), grade 1 (sum = 1), grade 2 (sum = 2–3), grade 3 (sum = 4–6), grade 4 (sum = 7–12). At the knee the osteophyte score was halved, due to four potential sites for osteophytes, before combining with joint-space narrowing grade to form the rOA score. The following thresholds were used for radiographic knee OA (rKOA): grade 0 (sum = 0), grade 1 (sum = 1), grade 2 (sum: >1–≤3), grade 3 (sum: >3–≤4.5) and grade 4 (sum: >4.5). When referencing rOA, unless otherwise stated, it refers to grade ≥2 OA at the specified joint(^10, 11^).

#### Demographic and clinical variables

BMI was derived from measures of participant weight and standing height. For both, measurements that coincided with the imaging visit were used. Race was determined using a self-reported questionnaire and when used as a covariate, was incorporated into the analyses as a binary variable (White vs. non-White), due to the low numbers of participants identifying as non-White. Although this variable was defined as ethnic background by UK Biobank, the categories better reflect racial groups. The Townsend Deprivation Index (TDI) was used as a proxy for social determinants of health. Higher scores of TDI, a postcode-based indicator of socioeconomic deprivation, signify greater deprivation(^16^). TDI was incorporated as a categorical variable based on pre-determined quintiles of deprivation(^17^) because UK Biobank is known to be less deprived than the general population. Self-reported chronic hip and knee pain were obtained through a questionnaire administered during the imaging visit.

Participants were asked two separate questions: “*Have you had [hip/knee] pains for more than 3 months?*” and a binary variable for both hip and knee joints was derived from their responses. Hospital diagnosed OA was derived from ICD-10 codes obtained from inpatient admissions, as previously published(^10, 11^).

#### Statistical analysis

Descriptive statistics were used to summarise the demographic characteristics, presented as means, standard deviations (SD) and ranges for continuous variables and as frequencies for categorical and binary variables. Pearson’s correlation coefficient (r) was used to compare minimum joint-space width at the hips and knees. Logistic regression was used to assess the associations between rOA grades 1-4 and self-reported joint pain. Similarly, logistic regression was applied to examine the interrelationships of rOA (grades ≥ 2), between bilateral joints (e.g., with OA in the left hip or knee modelled as the outcome and OA in the corresponding right joint as the exposure) and ipsilateral joints. These models were adjusted for age, sex, race, height, weight and TDI. Sex-stratified models were also run. Other models were used to examine associations between standardised BMI, height and weight with rHOA and rKOA. Logistic regression was used to assess associations of rOA at each joint (present vs absent) with TDI quintiles and race (White vs non-White). Results are reported as odds ratios (ORs) with 95% CIs. All statistical analyses were performed using Stata version 18 (StataCorp, College Station, TX, USA).

## Results

### Population characteristics

In total, 59,475 individuals (mean age: 65.1, SD: 7.7, range: 45-85 years), consisting of 31,414 females (52.8%), were included in this study. The mean height was 169.7 cm (range: 124-204 cm) and the mean body weight was 75.2 kg (range: 34-200 kg). The frequency of rHOA grade ≥2 was 4,098 (6.9%) for the right and 4,841 (8.1%) for the left hip. The corresponding frequency of rKOA grade ≥2 was 3,750 (6.3%) for the right and 4,220 (7.1%) for the left knee. Of those with features of rHOA, 1,285 (16.8%) had bilateral involvement. Likewise, of those with features of rKOA, 2,112 (36.1%) had bilateral involvement.

Among those with rOA affecting at least one joint, single-joint involvement was present in 8,745 (69.6%) individuals, two-joint involvement in 3,369 (26.8%) individuals, three-joint involvement in 394 (3.1%) individuals, and involvement in all four joints was found in 61 (0.5%) individuals. Of the 3,369 participants with two-joint rOA involvement, only 488 (14.5%) had one hip and one knee joint affected (i.e., non-cognate). Radiographic knee involvement was approximately twice as prevalent in females than in males, whereas radiographic hip involvement was twice as prevalent in males. Hospital diagnosed HOA was noted in 899 (1.5%) of individuals, while hospital diagnosed KOA was seen in 2,413 (4.1%) of individuals. Hip pain was self-reported in 4,924 individuals (8.3%), while knee pain was more common in 8,795 individuals (14.8%) (Table 1).

**Table 1.**
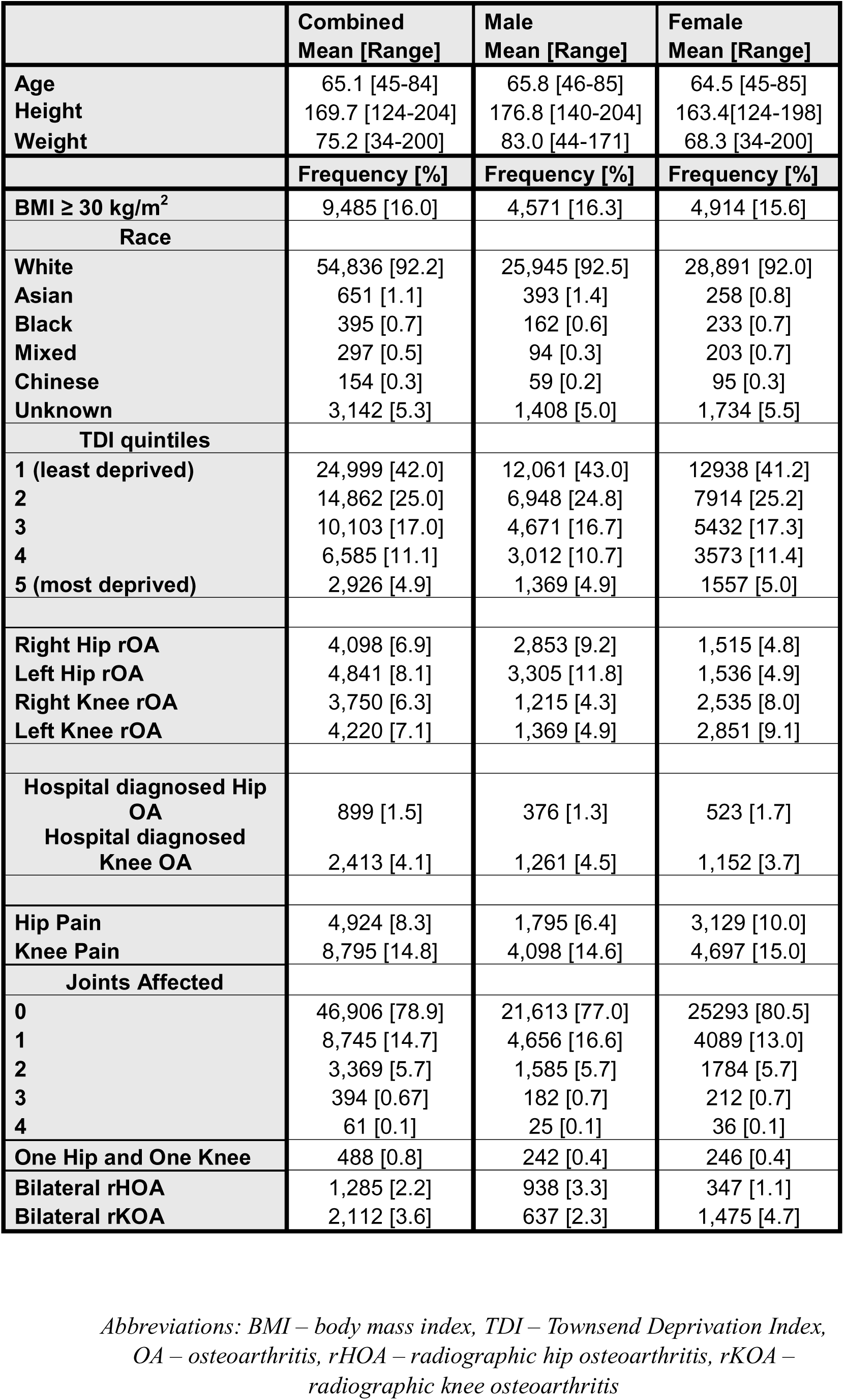
Descriptive statistics of study population.

|  | Combined<br>Mean [Range] | Male<br>Mean [Range] | Female<br>Mean [Range] |
| --- | --- | --- | --- |
| Age | 65.1 [45-84] | 65.8 [46-85] | 64.5 [45-85] |
| Height | 169.7 [124-204] | 176.8 [140-204] | 163.4 [124-198] |
| Weight | 75.2 [34-200] | 83.0 [44-171] | 68.3 [34-200] |
|  | Frequency [%] | Frequency [%] | Frequency [%] |
| BMI $\geq 30$ kg/m<sup>2</sup> | 9,485 [16.0] | 4,571 [16.3] | 4,914 [15.6] |
| Race |  |  |  |
| White | 54,836 [92.2] | 25,945 [92.5] | 28,891 [92.0] |
| Asian | 651 [1.1] | 393 [1.4] | 258 [0.8] |
| Black | 395 [0.7] | 162 [0.6] | 233 [0.7] |
| Mixed | 297 [0.5] | 94 [0.3] | 203 [0.7] |
| Chinese | 154 [0.3] | 59 [0.2] | 95 [0.3] |
| Unknown | 3,142 [5.3] | 1,408 [5.0] | 1,734 [5.5] |
| TDI quintiles |  |  |  |
| 1 (least deprived) | 24,999 [42.0] | 12,061 [43.0] | 12,938 [41.2] |
| 2 | 14,862 [25.0] | 6,948 [24.8] | 7,914 [25.2] |
| 3 | 10,103 [17.0] | 4,671 [16.7] | 5,432 [17.3] |
| 4 | 6,585 [11.1] | 3,012 [10.7] | 3,573 [11.4] |
| 5 (most deprived) | 2,926 [4.9] | 1,369 [4.9] | 1,557 [5.0] |
| Right Hip rOA | 4,098 [6.9] | 2,853 [9.2] | 1,515 [4.8] |
| Left Hip rOA | 4,841 [8.1] | 3,305 [11.8] | 1,536 [4.9] |
| Right Knee rOA | 3,750 [6.3] | 1,215 [4.3] | 2,535 [8.0] |
| Left Knee rOA | 4,220 [7.1] | 1,369 [4.9] | 2,851 [9.1] |
| Hospital diagnosed Hip OA | 899 [1.5] | 376 [1.3] | 523 [1.7] |
| Hospital diagnosed Knee OA | 2,413 [4.1] | 1,261 [4.5] | 1,152 [3.7] |
| Hip Pain | 4,924 [8.3] | 1,795 [6.4] | 3,129 [10.0] |
| Knee Pain | 8,795 [14.8] | 4,098 [14.6] | 4,697 [15.0] |
| Joints Affected |  |  |  |
| 0 | 46,906 [78.9] | 21,613 [77.0] | 25,293 [80.5] |
| 1 | 8,745 [14.7] | 4,656 [16.6] | 4,089 [13.0] |
| 2 | 3,369 [5.7] | 1,585 [5.7] | 1,784 [5.7] |
| 3 | 394 [0.67] | 182 [0.7] | 212 [0.7] |
| 4 | 61 [0.1] | 25 [0.1] | 36 [0.1] |
| One Hip and One Knee | 488 [0.8] | 242 [0.4] | 246 [0.4] |
| Bilateral rHOA | 1,285 [2.2] | 938 [3.3] | 347 [1.1] |
| Bilateral rKOA | 2,112 [3.6] | 637 [2.3] | 1,475 [4.7] |

Abbreviations: BMI – body mass index, TDI – Townsend Deprivation Index, OA – osteoarthritis, rHOA – radiographic hip osteoarthritis, rKOA – radiographic knee osteoarthritis

### Radiographic OA and associations with joint pain

Both the hip and knee joints demonstrated strong positive associations between grade of rOA and self-reported joint pain (Figure 2). Individuals with the most radiographically advanced changes (grade 4), had an 11- to 16-times increase in the odds of reporting chronic hip pain and 9- to 15-times increase in the odds of reporting chronic knee pain, as compared with those with no evidence of rOA. For those with minimal rKOA (grade 1), there was a 2-times increase in the odds of knee pain, while little evidence was seen for an association between grade 1 rHOA and hip pain (Supplementary Table 1). Across grades 1-3, the knee generally showed a stronger relationship with pain than the hip. Point estimates between contralateral joints were broadly similar but tended to be slightly higher for the right joint in most cases. Interestingly, rHOA showed moderate associations with knee pain, while rKOA showed no associations with hip pain (Supplementary Table 1).

**Figure 2.**
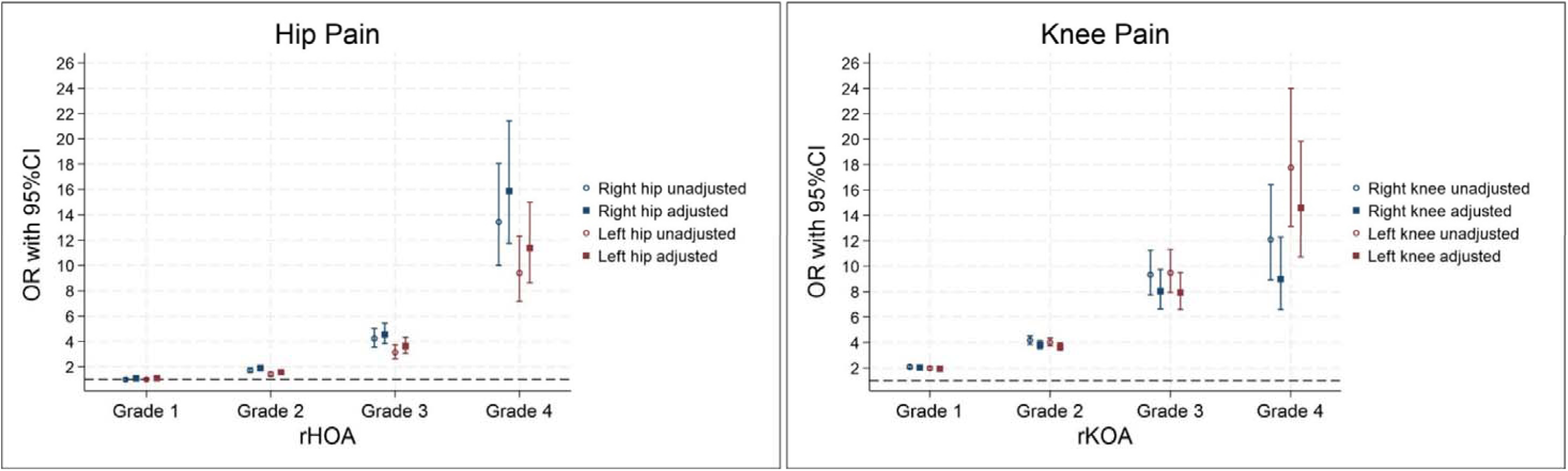
Logistic regression results for the associations between grades of radiographic osteoarthritis and self-reported joint pain Adjusted for age, sex, race, height, weight and deprivation rHOA – radiographic hip osteoarthritis, rKOA – radiographic knee osteoarthritis

In fully adjusted models, a greater number of joints affected by OA was associated with increased odds of experiencing hip and/or knee pain. Each additional OA joint exhibited higher odds of pain (Figure 3). Individuals with all four joints involved, had four times the odds of reporting pain compared with those without rOA (one joint: OR of 1.57 [95% CI 1.49-1.66], two joints: 2.58 [2.40-2.78], three joints: 3.22 [2.63-3.95], four joints: 4.29 [2.57-7.16]). When looking at hip and knee pain separately, similar positive associations were seen but with point estimates slightly higher for knee pain and lower for hip pain (Supplementary Figure 1).

**Figure 3.**
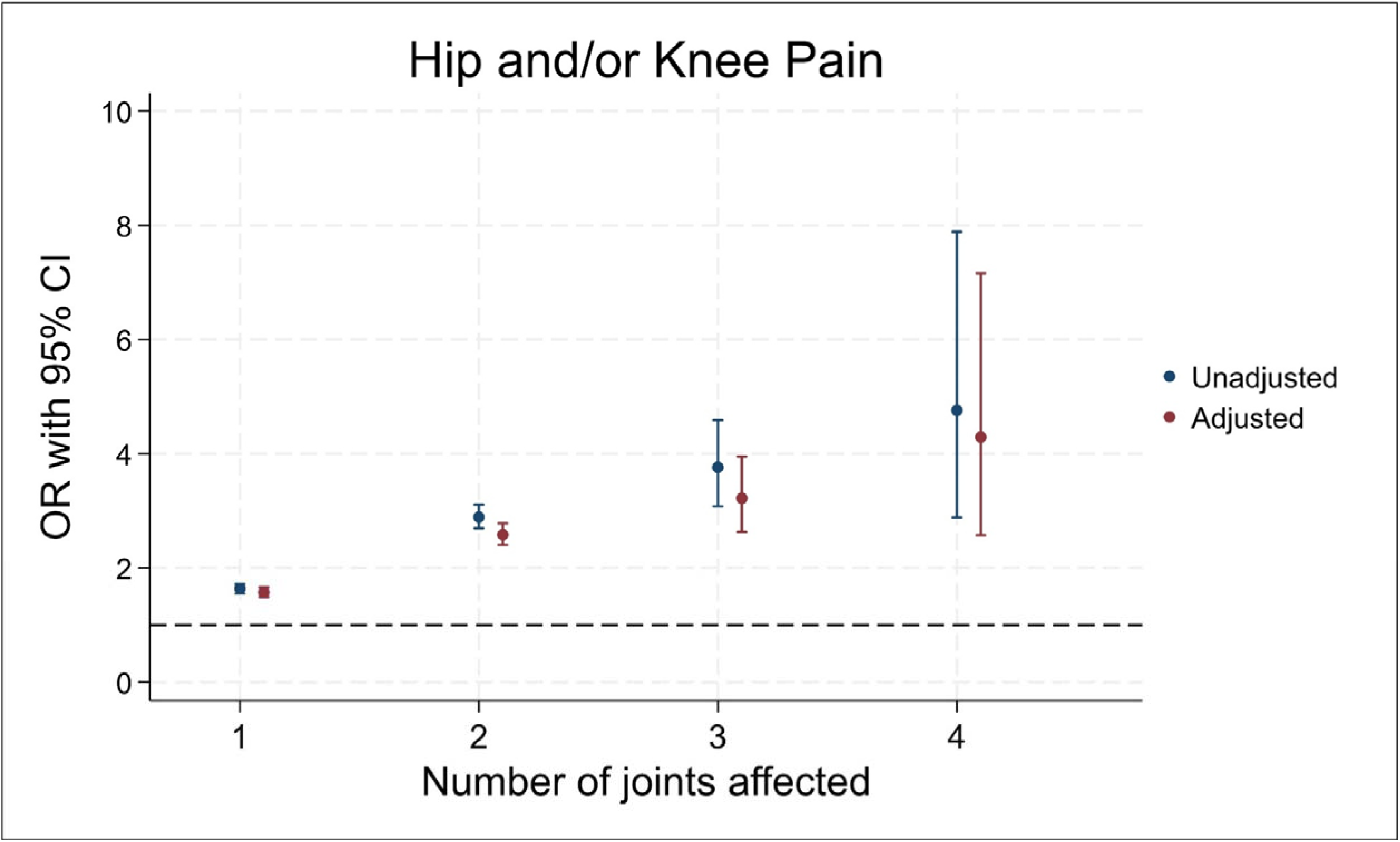
Logistic regression results for the associations between number of joints affected by radiographic osteoarthritis and self-reported hip and/or knee pain Adjusted for age, sex, race, height, weight and deprivation

### Joint site interrelationships

When comparing the odds of rOA between contralateral and ipsilateral hip and knee joints, both the hip and knee demonstrated robust bilateral associations (Figure 4). In fully-adjusted models, rHOA in one hip was associated with more than five times higher odds of rHOA in the other hip. Associations at the knees were much larger, with more than a 26-fold increase in the odds of right or left rKOA if the other joint was affected. Cross-joint associations, although considerably weaker, were still present: for individuals with right rHOA, the adjusted odds ratios were 1.25 (95% CI 1.10–1.41) for right rKOA and 1.36 (1.22–1.53) for left rKOA. Similar cross-joint (non-cognate) associations were observed for rHOA when rKOA was present. In sex stratified analyses, males showed stronger bilateral associations at the knees, while females showed stronger bilateral associations at the hip (Supplementary Table 2).

**Figure 4.**
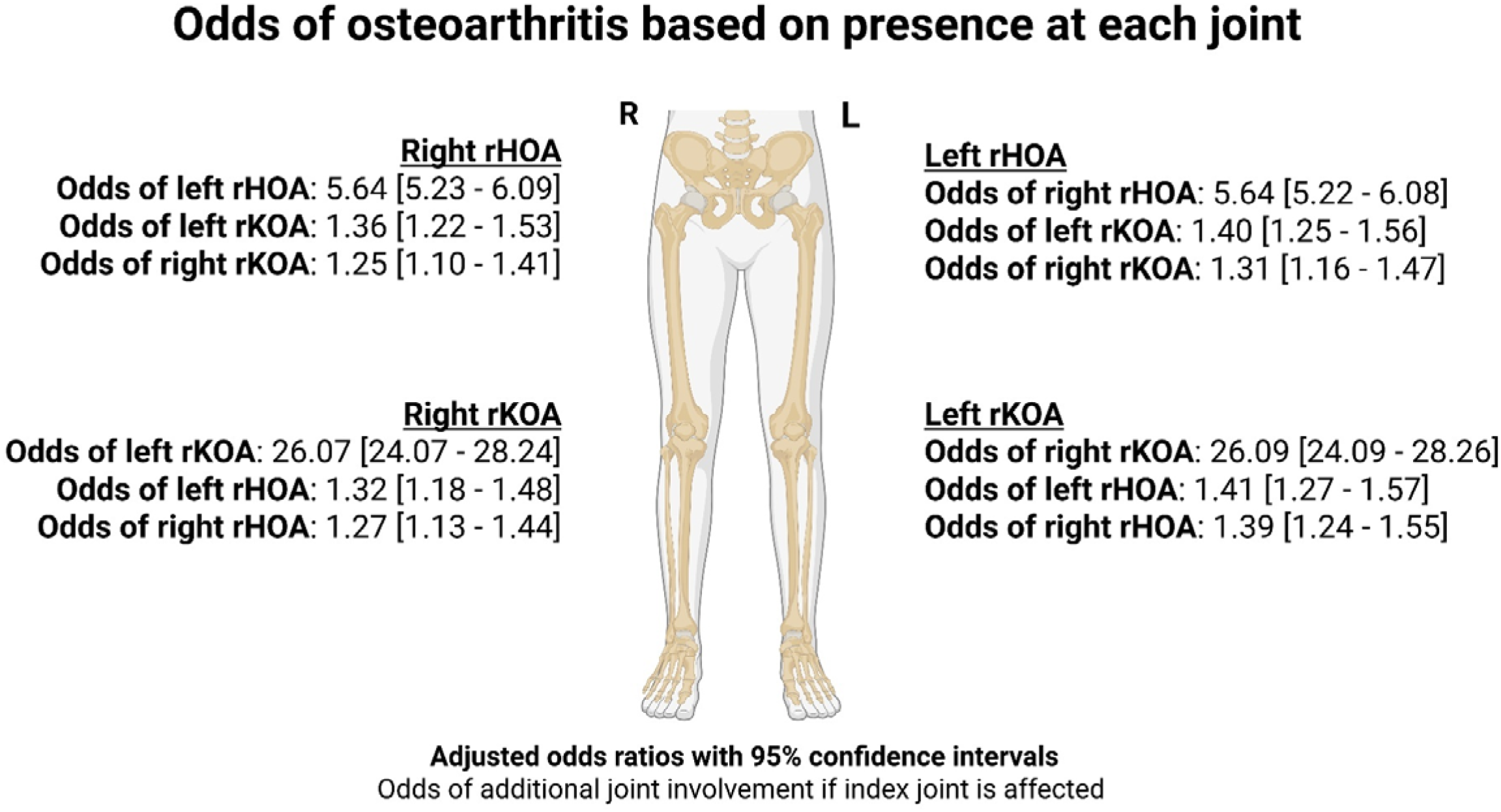
Logistic regression results for the odds of radiographic osteoarthritis based on presence at each joint Adjusted for age, sex, race, height, weight and deprivation rHOA – radiographic hip osteoarthritis, rKOA – radiographic knee osteoarthritis

Left and right minimum joint-space widths at the medial knee compartments showed a strong positive correlation with each other (r=0.73, p<0.001), while at the hips the positive correlation was slightly weaker (r=0.66, p<0.001). In comparison, the different anatomical joints demonstrated weaker correlations (r∼0.27-0.30, p<0.001) (Supplementary Figure 2).

### Associations with BMI, height and weight

BMI was strongly associated with rKOA (grades ≥2), with fully adjusted odds ratios per one standard deviation increase of 1.57 (95% CI 1.52–1.61) and 1.58 (1.53–1.62) for the right and left knees, respectively. In contrast, BMI was weakly associated with rHOA, with corresponding odds ratios of 1.05 (1.02–1.09) and 1.07 (1.03–1.10), for the right and left hips, respectively. Associations with obesity (BMI ≥30 kg/m^2^) showed a similar pattern, with the magnitude of association more than twice as large for the knees (right: OR 2.45 [2.27-2.65], left: OR 2.44 [2.27-2.62]), compared with the hips (right: OR 1.14 [1.05 - 1.24], left: OR 1.15 [1.06 - 1.24]). Likewise, rKOA showed stronger associations with weight. Greater height was associated with higher risk of rHOA (right: 1.33 [1.26-1.39], left: 1.27 [1.21 - 1.33]), but was inversely associated with lower risk of rKOA (right: OR 0.82 [0.77- 0.86], left: OR 0.83 [0.79 - 0.87] (Table 2).

**Table 2.**
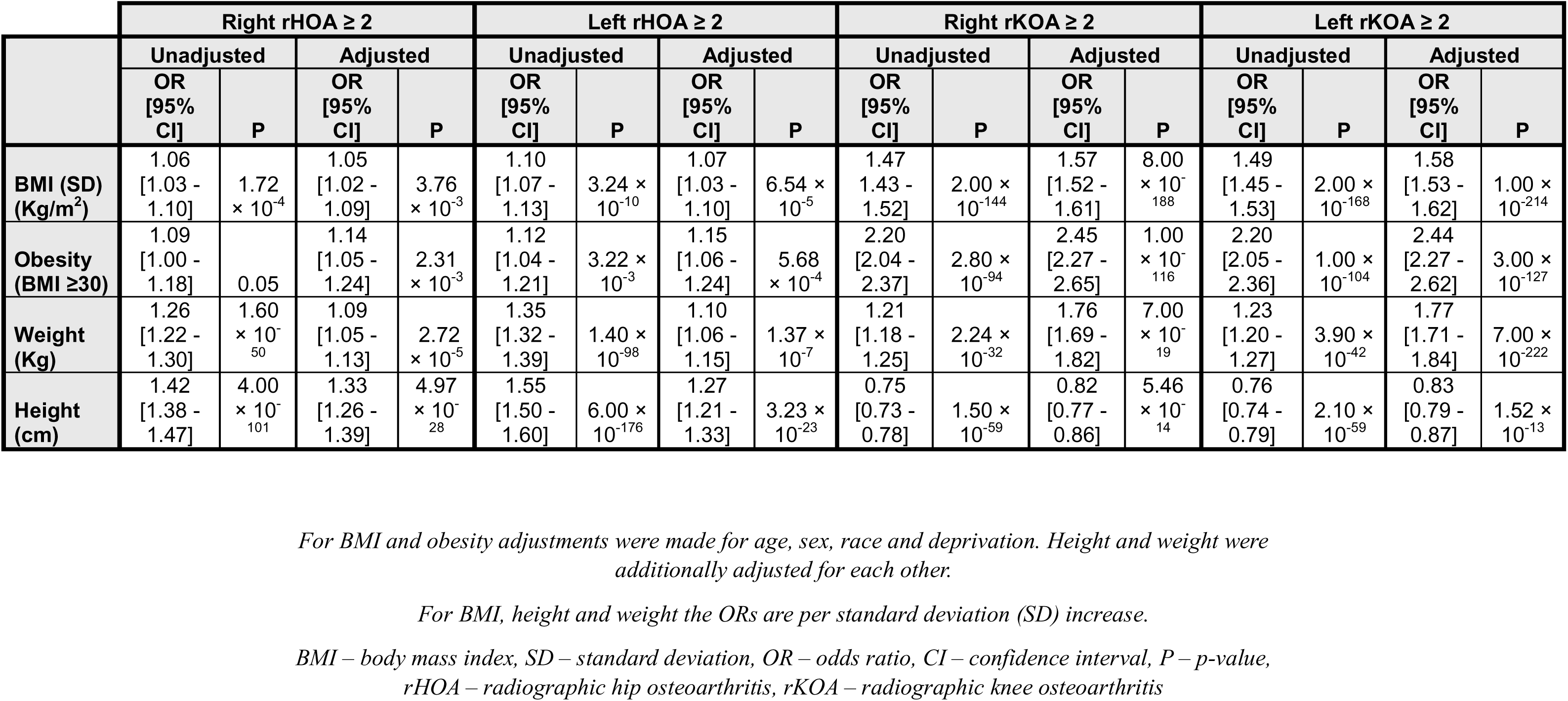
Logistic regression results for the associations between radiographic osteoarthritis and BMI, obesity, height and weight.

|  | Right rHOA ≥ 2 |  |  |  | Left rHOA ≥ 2 |  |  |  | Right rKOA ≥ 2 |  |  |  | Left rKOA ≥ 2 |  |  |  |
| --- | --- | --- | --- | --- | --- | --- | --- | --- | --- | --- | --- | --- | --- | --- | --- | --- |
|  | Unadjusted |  | Adjusted |  | Unadjusted |  | Adjusted |  | Unadjusted |  | Adjusted |  | Unadjusted |  | Adjusted |  |
|  | OR<br>[95%<br>CI] | P | OR<br>[95%<br>CI] | P | OR<br>[95%<br>CI] | P | OR<br>[95%<br>CI] | P | OR<br>[95%<br>CI] | P | OR<br>[95%<br>CI] | P | OR<br>[95%<br>CI] | P | OR<br>[95%<br>CI] | P |
| **BMI (SD)**<br>**(Kg/m<sup>2</sup>)** | 1.06<br>[1.03 -<br>1.10] | 1.72<br>× 10<sup>-4</sup> | 1.05<br>[1.02 -<br>1.09] | 3.76<br>× 10<sup>-3</sup> | 1.10<br>[1.07 -<br>1.13] | 3.24 ×<br>10<sup>-10</sup> | 1.07<br>[1.03 -<br>1.10] | 6.54 ×<br>10<sup>-5</sup> | 1.47<br>[1.43 -<br>1.52] | 2.00 ×<br>10<sup>-144</sup> | 1.57<br>[1.52 -<br>1.61] | 8.00<br>× 10<sup>-188</sup> | 1.49<br>[1.45 -<br>1.53] | 2.00 ×<br>10<sup>-168</sup> | 1.58<br>[1.53 -<br>1.62] | 1.00 ×<br>10<sup>-214</sup> |
| **Obesity**<br>**(BMI ≥30)** | 1.09<br>[1.00 -<br>1.18] | 0.05 | 1.14<br>[1.05 -<br>1.24] | 2.31<br>× 10<sup>-3</sup> | 1.12<br>[1.04 -<br>1.21] | 3.22 ×<br>10<sup>-3</sup> | 1.15<br>[1.06 -<br>1.24] | 5.68<br>× 10<sup>-4</sup> | 2.20<br>[2.04 -<br>2.37] | 2.80 ×<br>10<sup>-94</sup> | 2.45<br>[2.27 -<br>2.65] | 1.00<br>× 10<sup>-116</sup> | 2.20<br>[2.05 -<br>2.36] | 1.00 ×<br>10<sup>-104</sup> | 2.44<br>[2.27 -<br>2.62] | 3.00 ×<br>10<sup>-127</sup> |
| **Weight**<br>**(Kg)** | 1.26<br>[1.22 -<br>1.30] | 1.60<br>× 10<sup>-50</sup> | 1.09<br>[1.05 -<br>1.13] | 2.72<br>× 10<sup>-5</sup> | 1.35<br>[1.32 -<br>1.39] | 1.40 ×<br>10<sup>-98</sup> | 1.10<br>[1.06 -<br>1.15] | 1.37 ×<br>10<sup>-7</sup> | 1.21<br>[1.18 -<br>1.25] | 2.24 ×<br>10<sup>-32</sup> | 1.76<br>[1.69 -<br>1.82] | 7.00<br>× 10<sup>-19</sup> | 1.23<br>[1.20 -<br>1.27] | 3.90 ×<br>10<sup>-42</sup> | 1.77<br>[1.71 -<br>1.84] | 7.00 ×<br>10<sup>-222</sup> |
| **Height**<br>**(cm)** | 1.42<br>[1.38 -<br>1.47] | 4.00<br>× 10<sup>-101</sup> | 1.33<br>[1.26 -<br>1.39] | 4.97<br>× 10<sup>-28</sup> | 1.55<br>[1.50 -<br>1.60] | 6.00 ×<br>10<sup>-176</sup> | 1.27<br>[1.21 -<br>1.33] | 3.23 ×<br>10<sup>-23</sup> | 0.75<br>[0.73 -<br>0.78] | 1.50 ×<br>10<sup>-59</sup> | 0.82<br>[0.77 -<br>0.86] | 5.46<br>× 10<sup>-14</sup> | 0.76<br>[0.74 -<br>0.79] | 2.10 ×<br>10<sup>-59</sup> | 0.83<br>[0.79 -<br>0.87] | 1.52 ×<br>10<sup>-13</sup> |

For BMI and obesity adjustments were made for age, sex, race and deprivation. Height and weight were additionally adjusted for each other.
For BMI, height and weight the ORs are per standard deviation (SD) increase.
BMI – body mass index, SD – standard deviation, OR – odds ratio, CI – confidence interval, P – p-value, rHOA – radiographic hip osteoarthritis, rKOA – radiographic knee osteoarthritis

### Associations with deprivation and race

Prevalence of rOA was similar across different quintiles of deprivation and ethnic groups (Supplementary Table 3). Overall, deprivation (TDI quintiles) showed no consistent association with rOA at the hips or knees. For right rHOA, quintile 2 showed a slightly lower odds compared to quintile 1, the least deprived quintile, with an OR of 0.92 (0.85-1.00) Quintiles 3-5 showed no association with right rHOA (Supplementary Table 4). For left rHOA, the results were similarly null, as were those for both right and left rKOA. Equally, there was little evidence of consistent associations between rHOA and the different racial backgrounds. However, there was some evidence that Asian race is a risk factor for rKOA (right: 1.65 [1.20 - 2.26], left: 1.55 [1.14 - 2.10]) (Supplementary Table 5).

## Discussion

This large cohort study examined rOA of the hips and knees, to characterise joint interrelationships and describe associations with sex, pain, body size and deprivation. Our findings highlight differences between rHOA and rKOA. Overall, at each anatomical joint, increasing radiographic severity was associated with increasingly higher odds of self-reported joint pain. Likewise, a greater number of joints affected also saw increasing associations with pain. Notably, almost a third of patients with rOA experienced multi-joint involvement. Pronounced bilateral associations were observed, particularly at the knee but also at the hip, while cross-joint associations between the hip and knee, despite being weaker, were still present. Established risk factors for OA demonstrated clear joint-specific patterns, with BMI and weight being more strongly associated with rKOA than rHOA. Contrastingly, height was more strongly associated with rHOA but appeared protective in rKOA.

A notable finding of this study is the clinical relevance of DXA-derived rOA, with increasing severity associated with progressively higher odds of chronic joint pain, replicating smaller studies in UK Biobank(^10, 11^). These mirror associations seen in other cohorts, where increasing Kellgren and Lawrence (K/L) grade was associated with even greater odds of consistent knee pain(^18^). In MOST, K/L grade 4 was associated with 151 (43-526) times higher odds of frequent knee pain compared to grade 0. The corresponding odds ratio in the Framingham cohort was 73 (16.2-331). Presumably, these positive associations with symptoms reflect the fact that DXA-derived OA captures structurally relevant aspects of disease. Consistent with this suggestion, these measures have also been reported to be predictive of joint replacement(^10, 11^).

Several joint-specific differences were observed. In general rKOA showed stronger associations with knee pain than rHOA with hip pain. In particular, minimal rKOA (grade 1) was associated with increased odds of knee pain, no such association was seen for grade 1 rHOA. Previous research has shown that patients with early radiographic knee changes can experience pain and functional impairment similar to those with end-stage structural changes, while hip pain may only be strongly associated with more severely degraded joints (K/L ≥3)(^19, 20^). A cross-sectional study found 26% of osteoarthritic knee joints (KL ≥2) but only 7% of osteoarthritic hip joints were symptomatic(^21^). These differences could reflect the unique biomechanics of each joint. For example, the knee bears greater load and may be more sensitive to subtle changes caused by early disease(^22^). Nevertheless, our results and others show robust associations with hip pain in those with rHOA ≥2, reinforcing the clinical importance of radiographic changes at the hip(^23^).

Interestingly, rHOA showed modest associations with knee pain, while rKOA did not demonstrate a reciprocal association with hip pain. This aligns with existing understanding that pain originating from the hip is often referred to regions including the knee(^24^). A further consideration is the slightly stronger association between rOA and pain in right-sided joints as compared to the left. Most individuals are estimated to be right-footed so this difference may reflect leg dominance(^25^). Preferential use of the dominant limb may result in greater cumulative mechanical loading, which leads to greater symptom manifestation from the structural abnormality, noting the frequency of rOA is not increased on the right side.

This study provides updated frequency estimates for rOA in a large general cohort. The slightly lower occurrence of rKOA compared to rHOA, contrasts with previous studies, including the Johnston County OA Project and the Rotterdam Study, which generally report a higher prevalence of knee than hip OA(^26–28^). Similarly, recent UK-based clinical estimates suggest that approximately 5.4 million people are affected by knee OA, compared with 3.2 million with hip OA(^29^). Given that the prevalence of pain and hospital diagnosed OA at the knee in UK Biobank was double that of hip, this discrepancy likely reflects differences in how early radiographic change presents with symptoms between the joints. Furthermore, it is difficult to make grading systems that are perfectly aligned. For example, our grading approach may have favoured the detection of hip disease, where more weight was given to the presence of osteophytes. The observed sex-differences in the prevalence of rHOA and rKOA are consistent with previous estimates, with rKOA more common among females and rHOA more common among males(^30^). These differences may be due to variations in joint shape, hormonal factors and pain sensitivity, but further research is required to fully understand the causes. That said, it is important for clinicians to be aware of this sex-based variation in prevalence.

Radiographic OA in one knee markedly increased the likelihood of involvement in the contralateral knee. Similarly, robust bilateral associations were observed at the hip joints. Supporting this finding, a previous study found that 80% of participants with unilateral rKOA progressed to bilateral disease over 12 years(^31^), while in another study 40% of patients with unilateral rHOA developed bilateral rHOA over 9 years(^32^). In contrast, cross-joint associations between the hip and knee were considerably weaker, consistent with an earlier finding that previous hip OA is only modestly associated with incident knee OA (Hazard ratio (HR) of 1.36) and vice versa for knee OA and incident hip OA (HR 1.66)(^33^). These findings support studies suggesting the genetic architecture of OA is joint-specific, with distinct loci contributing towards disease risk at different anatomical sites(^34^). For example, the largest genome wide association study to date, found 151 independent associations for hip and 146 for knee OA(^35^).

Also, minimum joint-space width, which is known to be genetically determined(^36^), was strongly correlated when comparing bilateral hips and knees but only weakly correlated across joint types. Environmental factors, such as abnormal compensatory loading patterns in the contralateral knees of patients with unilateral knee OA, are also of likely importance(^37^).

While shared systemic factors may contribute to lower limb OA across other joint types, clinicians should be aware that further joint involvement appears to preferentially occur by anatomical site. This may help guide monitoring strategies and the development of targeted treatment to prevent multi-joint progression.

Consistent with this, we observed different associations between common anthropometric risk factors and rOA at the hip and knee. BMI, obesity and weight were strongly associated with rKOA but showed weak associations with rHOA. Height displayed an opposing association, being positively associated with rHOA and negatively associated with rKOA. Research has established that high BMI is more strongly associated with knee than hip OA(^38^), which likely reflects anatomical and pathophysiological differences with the knees being more vulnerable to increased mechanical load and obesity-induced chronic inflammation(^22, 39^). Previously it has also been demonstrated how the knee and hips vary in terms of susceptibility to occupational risk factors such as shift work and heavy manual work(^40^). These different risk factor profiles suggest that joint-specific treatment strategies should be considered. Indeed, a recent randomised controlled trial has shown that treatment with the GLP-1 agonist semaglutide, a weight-loss injectable, can significantly reduce pain in knee OA patients(^41^). These findings have been corroborated in pre-clinical studies(^42^). Our results, and those of others, indicate that such strategies may prove less effective in hip OA.

The association between hip OA and height suggests shared mechanisms with growth. Genetic studies have identified loci associated with hip OA, height and cartilage thickness, supporting a role for growth-related pathways in the development of hip disease(^36, 43^). Height-associated loci have also been related to skeletal development and bone morphology(^44^), which may be considered as risk factors for HOA. Evidence regarding height and its association with knee OA remains conflicting, with studies identifying taller stature to be associated with both increased and decreased risk(^45, 46^).

In this UK Biobank study, the majority of those with multi-joint OA were bilateral knee or hip OA suggesting they are distinct conditions. Though, a minority of individuals with OA (7.5%) exist with non-cognate multi-joint disease (e.g., co-existent hip and knee OA), where the likelihood of reporting hip and/or knee pain increased with each additional joint affected. These individuals represent a more generalized pattern of disease(^47^). Unfortunately, imaging is not available in UK Biobank to derive radiographic hand OA and identify associations with lower limb rOA. Nevertheless, the involvement of multiple weight-bearing joints has previously been associated with greater pain severity, poorer functional outcomes and increased risk of falls(^5, 48, 49^). Together, these findings highlight the clinical importance of multi-joint OA as a concept and suggest those patients with higher disease burden merit the consideration of cumulative joint involvement, rather than isolated single-joint pathology.

Although higher incidence of OA has previously been reported among more deprived populations(^3^), we observed little association between deprivation quintiles and rOA at either joint site. Similarly, there was no strong evidence that race was a risk factor for rHOA. However, Asian race was a risk factor for rKOA. Nonetheless, these analyses were limited by the low numbers of participants with rOA among racial minorities and a greater skew towards less deprived individuals. Notably, inequalities have previously been identified in the provision of joint replacement, with rates lower among less affluent areas, despite greater clinical need(^50^). Disparities in access to care exist and this study underscores the need to ensure equitable access across all quintiles of deprivation.

A key strength of this study is its large sample size, providing substantial statistical power to these analyses. The use of image-derived radiographic measures has enabled detailed assessment of disease severity and for direct comparison of the hip and knee. Despite this, the study is limited by its cross-sectional nature, preventing the deduction of causal associations. Another limitation is the lack of ethnic or socioeconomic diversity among the UK Biobank population, limiting generalisability to other populations. Finally, this study is based on supine DXAs and are therefore not weight-bearing. That said, the replicated strong clinical associations support their use for deriving rOA.

In conclusion, this large cross-sectional study demonstrates that rOA captures structurally and clinically meaningful disease, while also identifying important heterogeneity between hip and knee OA. Increasing radiographic severity and multi-joint involvement were increasingly associated with pain. Strong bilateral but weaker cross-joint associations, alongside unique relationships with BMI, height, and weight, support the existence of different phenotypes. These findings indicate the importance of tailored approaches in the monitoring and management of OA. Further research is required to explore joint site interrelationships over time and to inform interventions aimed at preventing or managing multi-joint OA. Additionally, research is warranted to elucidate the joint-specific pathomechanisms.

## Acknowledgements and affiliations

The authors would like to acknowledge the huge contribution of Dr Monika Frysz and Dr Rhona Beynon in establishing this unprecedented imaging resource in UK Biobank. Both individuals declined the offer of authorship as they have left academia.

## Data statement

All UK Biobank data (project number 17295) will be available via their data showcase shortly after publication.

## References

1. Hunter DJ, Bierma-Zeinstra S. Osteoarthritis. The Lancet. 2019;393(10182):1745–59.

2. Global, regional, and national burden of osteoarthritis, 1990-2020 and projections to 2050: a systematic analysis for the Global Burden of Disease Study 2021. Lancet Rheumatol. 2023;5(9):e508–e22.

3. Reyes C, Garcia-Gil M, Elorza JM, Mendez-Boo L, Hermosilla E, Javaid MK, et al. Socio-economic status and the risk of developing hand, hip or knee osteoarthritis: a region-wide ecological study. Osteoarthritis and Cartilage. 2015;23(8):1323–9.

4. Abdullah Y, Olubowale OO, Hackshaw KV. Racial disparities in osteoarthritis: Prevalence, presentation, and management in the United States. Journal of the National Medical Association. 2025;117(1):55–60.

5. Nelson AE. Multiple joint osteoarthritis (MJOA): What’s in a name? Osteoarthritis Cartilage. 2024;32(3):234–40.

6. Collins KH, Haugen IK, Neogi T, Guilak F. Osteoarthritis as a systemic disease. Nature Reviews Rheumatology. 2026;22(2):105–17.

7. Boer CG, Hatzikotoulas K, Southam L, Stefánsdóttir L, Zhang Y, Coutinho de Almeida R, et al. Deciphering osteoarthritis genetics across 826,690 individuals from 9 populations. Cell. 2021;184(18):4784–818.e17.

8. Hatzikotoulas K, Southam L, Stefansdottir L, Boer CG, McDonald M-L, Pett JP, et al. Translational genomics of osteoarthritis in 1,962,069 individuals. Nature. 2025;641(8065):1217–24.

9. Littlejohns TJ, Holliday J, Gibson LM, Garratt S, Oesingmann N, Alfaro-Almagro F, et al. The UK Biobank imaging enhancement of 100,000 participants:[rationale, data collection, management and future directions. Nat Commun. 2020;11(1):2624.

10. Beynon RA, Saunders FR, Ebsim R, Faber BG, Jung M, Gregory JS, et al. A novel classifier of radiographic knee osteoarthritis for use on knee DXA images is predictive of joint replacement in UK Biobank. Rheumatology Advances in Practice. 2025;9(1).

11. Faber BG, Ebsim R, Saunders FR, Frysz M, Lindner C, Gregory JS, et al. A novel semi-automated classifier of hip osteoarthritis on DXA images shows expected relationships with clinical outcomes in UK Biobank. Rheumatology (Oxford). 2022;61(9):3586–95.

12. Bycroft C, Freeman C, Petkova D, Band G, Elliott LT, Sharp K, et al. The UK Biobank resource with deep phenotyping and genomic data. Nature. 2018;562(7726):203–9.

13. Littlejohns TJ, Holliday J, Gibson LM, Garratt S, Oesingmann N, Alfaro-Almagro F, et al. The UK Biobank imaging enhancement of 100,000 participants:[rationale, data collection, management and future directions. Nature Communications. 2020;11(1):2624.

14. Faber BG, Ebsim R, Saunders FR, Frysz M, Lindner C, Gregory JS, et al. Osteophyte size and location on hip DXA scans are associated with hip pain: findings from a cross sectional study in UK Biobank. Bone. 2021:116146.

15. Beynon RA, Saunders FR, Ebsim R, Faber BG, Jung M, Gregory JS, et al. A novel classifier of radiographic knee osteoarthritis for use on knee DXA images is predictive of joint replacement in UK Biobank. Rheumatol Adv Pract. 2025;9(1):rkaf009.

16. Townsend P, Phillimore, P., & Beattie, A. Health and Deprivation: Inequality and the North. London: Routledge; 1988. 236 p.

17. Hippisley-Cox J. Cut offs for Townsend quintiles in England and Wales and QResearch 2011 [23/11/25]. Available from: https://www.qresearch.org/media/1056/cut-offs-for-townsend-quintiles-in-england-and-wales-and-qresearch.pdf.

18. Neogi T, Felson D, Niu J, Nevitt M, Lewis CE, Aliabadi P, et al. Association between radiographic features of knee osteoarthritis and pain: results from two cohort studies. Bmj. 2009;339:b2844.

19. Jones LD, Bottomley N, Harris K, Jackson W, Price AJ, Beard DJ. The clinical symptom profile of early radiographic knee arthritis: a pain and function comparison with advanced disease. Knee Surgery, Sports Traumatology, Arthroscopy. 2016;24(1):1795.

20. Iidaka T, Muraki S, Akune T, Oka H, Kodama R, Tanaka S, et al. Prevalence of radiographic hip osteoarthritis and its association with hip pain in Japanese men and women: the ROAD study. Osteoarthritis Cartilage. 2016;24(1):117–23.

21. Pereira D, Severo M, Santos RA, Barros H, Branco J, Lucas R, et al. Knee and hip radiographic osteoarthritis features: differences on pain, function and quality of life. Clinical Rheumatology. 2016;35(6):1555–64.

22. Hall M, van der Esch M, Hinman RS, Peat G, de Zwart A, Quicke JG, et al. How does hip osteoarthritis differ from knee osteoarthritis? Osteoarthritis and Cartilage. 2022;30(1):32–41.

23. Rondas GA, Macri EM, Oei EH, Bierma-Zeinstra SM, Rijkels-Otters HB, Runhaar J. Association between hip pain and radiographic hip osteoarthritis in primary care: the CHECK cohort. Br J Gen Pract. 2022;72(723):e722–8.

24. Dibra FF, Prieto HA, Gray CF, Parvataneni HK. Don’t forget the hip! Hip arthritis masquerading as knee pain. Arthroplast Today. 2018;4(1):118–24.

25. Tran US, Voracek M. Footedness Is Associated with Self-reported Sporting Performance and Motor Abilities in the General Population. Front Psychol. 2016;7:1199.

26. Odding E, Valkenburg HA, Algra D, Vandenouweland FA, Grobbee DE, Hofman A. Associations of radiological osteoarthritis of the hip and knee with locomotor disability in the Rotterdam Study. Ann Rheum Dis. 1998;57(4):203–8.

27. Nelson AE, Hu D, Arbeeva L, Alvarez C, Cleveland RJ, Schwartz TA, et al. Point prevalence of hip symptoms, radiographic, and symptomatic OA at five time points: The Johnston County Osteoarthritis Project, 1991–2018. Osteoarthritis and Cartilage Open. 2022;4(2):100251.

28. Nelson AE, Hu D, Arbeeva L, Alvarez C, Cleveland RJ, Schwartz TA, et al. The Prevalence of Knee Symptoms, Radiographic, and Symptomatic Osteoarthritis at Four Time Points: The Johnston County Osteoarthritis Project, 1999-2018. ACR Open Rheumatol. 2021;3(8):558–65.

29. Arthritis UK. The State of Musculoskeletal Health 2025. 2025. p. 41.

30. Faber BG, Macrae F, Jung M, Zucker BE, Beynon RA, Tobias JH. Sex differences in the radiographic and symptomatic prevalence of knee and hip osteoarthritis. Front Endocrinol (Lausanne). 2024;15:1445468.

31. Metcalfe AJ, Andersson ML, Goodfellow R, Thorstensson CA. Is knee osteoarthritis a symmetrical disease? Analysis of a 12 year prospective cohort study. BMC Musculoskelet Disord. 2012;13:153.

32. Vossinakis IC, Georgiades G, Kafidas D, Hartofilakidis G. Unilateral hip osteoarthritis: can we predict the outcome of the other hip? Skeletal Radiology. 2008;37(10):911–6.

33. Prieto-Alhambra D, Judge A, Javaid MK, Cooper C, Diez-Perez A, Arden NK. Incidence and risk factors for clinically diagnosed knee, hip and hand osteoarthritis: influences of age, gender and osteoarthritis affecting other joints. Ann Rheum Dis. 2014;73(9):1659–64.

34. Zhai G, Huang J. Genetics of osteoarthritis. Best Practice & Research Clinical Rheumatology. 2024;38(4):101972.

35. Hatzikotoulas K, Southam L, Stefansdottir L, Boer CG, McDonald M-L, Pett JP, et al. Translational genomics of osteoarthritis in 1,962,069 individuals. Nature. 2025;641(8065):1217–24.

36. Faber BG, Frysz M, Boer CG, Evans DS, Ebsim R, Flynn KA, et al. The identification of distinct protective and susceptibility mechanisms for hip osteoarthritis: findings from a genome-wide association study meta-analysis of minimum joint space width and Mendelian randomisation cluster analyses. EBioMedicine. 2023;95:104759.

37. Metcalfe A, Stewart C, Postans N, Dodds A, Smith H, Holt C, et al. BIOMECHANICS OF THE UNAFFECTED JOINTS IN PATIENTS WITH KNEE OSTEOARTHRITIS. Orthopaedic Proceedings. 2012;94-B(SUPP_XVIII):41-.

38. Reyes C, Leyland KM, Peat G, Cooper C, Arden NK, Prieto-Alhambra D. Association Between Overweight and Obesity and Risk of Clinically Diagnosed Knee, Hip, and Hand Osteoarthritis: A Population-Based Cohort Study. Arthritis Rheumatol. 2016;68(8):1869–75.

39. Shumnalieva R, Kotov G, Ermencheva P, Monov S. Pathogenic Mechanisms and Therapeutic Approaches in Obesity-Related Knee Osteoarthritis. Biomedicines. 2023;12(1).

40. Hashmi A, Scott S, Jung M, Meng QJ, Tobias JH, Beynon RA, et al. Associations between work characteristics and osteoarthritis: A cross-sectional study of 285,947 UK Biobank participants. Osteoarthr Cartil Open. 2025;7(1):100565.

41. Bliddal H, Bays H, Czernichow S, Hemmingsson JU, Hjelmesæth J, Morville TH, et al. Once-Weekly Semaglutide in Persons with Obesity and Knee Osteoarthritis. New England Journal of Medicine. 2024;391(17):1573–83.

42. Qin H, Yu J, Yu H, Zhou C, Yuan D, Wang Z, et al. Semaglutide ameliorates osteoarthritis progression through a weight loss-independent metabolic restoration mechanism. Cell Metab. 2026.

43. Aspden RM. Osteoarthritis: a problem of growth not decay? Rheumatology (Oxford). 2008;47(10):1452–60.

44. Styrkarsdottir U, Stefansson OA, Gunnarsdottir K, Thorleifsson G, Lund SH, Stefansdottir L, et al. GWAS of bone size yields twelve loci that also affect height, BMD, osteoarthritis or fractures. Nature Communications. 2019;10(1):2054.

45. Welling M, Auvinen J, Lehenkari P, Männikkö M, Karppinen J, Eskola PJ. Association between height and osteoarthritis of the knee and hip: The Northern Finland Birth Cohort 1966 Study. Int J Rheum Dis. 2017;20(9):1095–104.

46. Lee DH, Lee HS, Jang SH, Heu JY, Han K, Lee SW. Decreased Risk of Knee Osteoarthritis with Taller Height in an East Asian Population: A Nationwide Cohort Study. J Clin Med. 2023;13(1).

47. Finney A, Dziedzic KS, Lewis M, Healey E. Multisite peripheral joint pain: a cross-sectional study of prevalence and impact on general health, quality of life, pain intensity and consultation behaviour. BMC Musculoskelet Disord. 2017;18(1):535.

48. Pavel RMS, Purza AL, Tit DM, Radu A-F, Iovanovici DC, Vasileva D, et al. Functional Burden and Quality of Life in Hip and Knee Osteoarthritis: A Cross-Sectional Study. Medicina. 2025;61(7):1155.

49. Smith RD, McHugh GA, Quicke JG, Finney A, Lewis M, Dziedzic KS, et al. The relationship between multisite peripheral joint pain and physical activity levels in older adults: A cross-sectional survey. Musculoskeletal Care. 2022;20(2):341–8.

50. Lenguerrand E, Ben-Shlomo Y, Rangan A, Beswick A, Whitehouse MR, Deere K, et al. Inequalities in provision of hip and knee replacement surgery for osteoarthritis by age, sex, and social deprivation in England between 2007-2017: A population-based cohort study of the National Joint Registry. PLoS Med. 2023;20(4):e1004210.

