## Supplementary Tables and Figures for "A comprehensive descriptive analysis of hip and knee radiographic osteoarthritis in the UK Biobank in relation to joint pain, joint site interrelationships, obesity, race and deprivation: Findings from 59,475 individuals"

Supplementary Table 1 - Logistic regression results showing the associations between increasing grades of radiographic osteoarthritis and self-reported pain

|  |  | **Odds of hip Pain** | |  | **Odds of knee pain** | | | |
| --- | --- | --- | --- | --- | --- | --- | --- | --- |
|  | **Unadjusted** | | **Fully Adjusted** | | **Unadjusted** | | **Fully Adjusted** | |
|  | **OR [95% CI]** | **P** | **OR [95% CI]** | **P** | **OR [95% CI]** | **P** | **OR [95% CI]** | **P** |
| **Right rHOA 1** | 0.99 [0.90 - 1.09] | 0.79 | 1.08 [0.98 - 1.19] | 0.14 | 1.12 [1.04 - 1.20] | 1.57 × 10^-3^ | 1.16 [1.08 - 1.25] | 4.37 × 10^-5^ |
| **Right rHOA 2** | 1.72 [1.54 - 1.92] | 2.11 × 10^-22^ | 1.89 [1.69 - 2.11] | 9.32 × 10^-29^ | 1.06 [0.96 - 1.17] | 0.22 | 1.04 [0.94 - 1.15] | 0.44 |
| **Right rHOA 3** | 4.23 [3.55 - 5.03] | 2.50 × 10^-59^ | 4.55 [3.81 - 5.44] | 1.80 × 10^-62^ | 1.29 [1.06 - 1.57] | 0.01 | 1.25 [1.02 - 1.52] | 0.03 |
| **Right rHOA 4** | 13.44 [10.01 - 18.04] | 5.60 × 10^-67^ | 15.87 [11.75 - 21.43] | 1.40 × 10^-72^ | 1.57 [1.10 - 2.25] | 0.01 | 1.58 [1.10 - 2.27] | 0.01 |
| **Left rHOA 1** | 1.00 [0.91 - 1.10] | 0.98 | 1.09 [1.00 - 1.20] | 0.06 | 1.13 [1.06 - 1.21] | 4.32 × 10^-4^ | 1.16 [1.09 - 1.25] | 1.92 × 10^-5^ |
| **Left rHOA 2** | 1.41 [1.26 - 1.57] | 6.26 × 10^-10^ | 1.58 [1.41 - 1.76] | 1.10 × 10^-15^ | 1.28 [1.17 - 1.40] | 2.45 × 10^-8^ | 1.25 [1.14 - 1.37] | 9.63 × 10^-7^ |
| **Left rHOA 3** | 3.14 [2.64 - 3.73] | 1.02 × 10^-38^ | 3.63 [3.04 - 4.33] | 1.40 × 10^-46^ | 1.26 [1.05 - 1.51] | 0.01 | 1.25 [1.03 - 1.50] | 0.02 |
| **Left rHOA 4** | 9.40 [7.17 - 12.31] | 1.80 × 10^-59^ | 11.37 [8.63 - 14.99] | 7.60 × 10^-67^ | 1.94 [1.42 - 2.64] | 3.10 × 10^-5^ | 2.02 [1.47 - 2.76] | 1.32 × 10^-5^ |
| **Right rKOA 1** | 1.17 [1.07 - 1.27] | 3.47 × 10^-4^ | 0.96 [0.88 - 1.04] | 0.31 | 2.09 [1.96 - 2.22] | 2.00 × 10^-121^ | 2.04 [1.91 - 2.18] | 2.00 × 10^-105^ |
| **Right rKOA 2** | 1.27 [1.13 - 1.44] | 1.01 × 10^-4^ | 0.98 [0.86 - 1.11] | 0.73 | 4.17 [3.85 - 4.50] | 2.00 × 10^-279^ | 3.82 [3.52 - 4.15] | 4.00 × 10^-227^ |
| **Right rKOA 3** | 1.30 [0.95 - 1.76] | 0.10 | 0.93 [0.68 - 1.27] | 0.65 | 9.33 [7.74 - 11.26] | 5.00 × 10^-121^ | 8.04 [6.64 - 9.74] | 6.00 × 10^-101^ |
| **Right rKOA 4** | 0.98 [0.57 - 1.69] | 0.95 | 0.60 [0.34 - 1.04] | 0.07 | 12.10 [8.93 - 16.40] | 2.70 × 10^-58^ | 8.99 [6.58 - 12.28] | 3.10 × 10^-43^ |
| **Left rKOA 1** | 1.14 [1.05 - 1.24] | 1.19 × 10^-3^ | 0.94 [0.87 - 1.03] | 0.17 | 1.98 [1.87 - 2.11] | 6.00 × 10^-111^ | 1.93 [1.81 - 2.06] | 9.20 × 10^-95^ |
| **Left rKOA 2** | 1.22 [1.09 - 1.38] | 6.99 × 10^-4^ | 0.93 [0.82 - 1.04] | 0.21 | 4.02 [3.73 - 4.34] | 3.00 × 10^-289^ | 3.66 [3.39 - 3.96] | 5.00 × 10^-231^ |
| **Left rKOA 3** | 1.16 [0.86 - 1.58] | 0.33 | 0.80 [0.59 - 1.10] | 0.17 | 9.48 [7.93 - 11.33] | 2.00 × 10^-134^ | 7.92 [6.59 - 9.51] | 1.00 × 10^-108^ |
| **Left rKOA 4** | 1.32 [0.84 - 2.08] | 0.23 | 0.88 [0.56 - 1.40] | 0.60 | 17.76 [13.14 - 24.01] | 4.10 × 10^-78^ | 14.60 [10.74 - 19.83] | 7.80 × 10^-66^ |

*Adjusted for age, sex, race, height, weight and deprivation*

*rHOA – radiographic hip osteoarthritis, rKOA – radiographic knee osteoarthritis, OR – odds ratio, P – p-value*

Supplementary Table 2 - Logistic regression results for the odds of osteoarthritis based on presence at each joint in combined and sex-stratified analyses

|  | **COMBINED** | | | | **MALE** | | | | **FEMALE** | | | |
| --- | --- | --- | --- | --- | --- | --- | --- | --- | --- | --- | --- | --- |
|  | **Unadjusted** | | **Fully Adjusted** | | **Unadjusted** | | **Fully Adjusted** | | **Unadjusted** | | **Fully Adjusted** | |
|  | **OR [95% CI]** | **P** | **OR [95% CI]** | **P** | **OR [95% CI]** | **P** | **OR [95% CI]** | **P** | **OR [95% CI]** | **P** | **OR [95% CI]** | **P** |
| Right rHOA |  |  |  |  |  |  |  |  |  |  |  |  |
| **Odds of left hip rOA** | 6.66 [6.18 - 7.17] | <0.01 × 10^-324^ | 5.64 [5.23 - 6.09] | <0.01 × 10^-324^ | 5.57 [5.09 - 6.10] | 1.00 × 10^-301^ | 5.37 [4.90 - 5.88] | 4.00 × 10^-285^ | 7.17 [6.28 - 8.20] | 5.00 × 10^-185^ | 6.11 [5.33 - 7.00] | 8.00 × 10^-150^ |
| **Odds of right knee rOA** | 1.31 [1.17 - 1.48] | 7.11 × 10^-6^ | 1.25 [1.10 - 1.41] | 4.15 × 10^-4^ | 1.29 [1.08 - 1.55] | 5.99 × 10^-3^ | 1.16 [0.97 - 1.39] | 0.11 | 1.65 [1.41 - 1.94] | 5.70 × 10^-10^ | 1.32 [1.12 - 1.56] | 8.27 × 10^-4^ |
| **Odds of Left knee rOA** | 1.41 [1.26 - 1.57] | 1.04 × 10^-9^ | 1.36 [1.22 - 1.53] | 7.96 × 10^-8^ | 1.38 [1.16 - 1.63] | 1.98 × 10^-4^ | 1.25 [1.06 - 1.48] | 9.88 × 10^-3^ | 1.79 [1.55 - 2.08] | 1.02 × 10^-14^ | 1.47 [1.26 - 1.71] | 8.78 × 10^-7^ |
| Left rHOA |  |  |  |  |  |  |  |  |  |  |  |  |
| **Odds of right hip rOA** | 6.66 [6.18 - 7.17] | <0.01 × 10^-324^ | 5.64 [5.22 - 6.08] | <0.01 × 10^-324^ | 5.57 [5.09 - 6.10] | 1.00 × 10^-301^ | 5.37 [4.90 - 5.88] | 3.00 × 10^-285^ | 7.17 [6.28 - 8.20] | 5.00 × 10^-185^ | 6.12 [5.34 - 7.01] | 8.00 × 10^-150^ |
| **Odds of right knee rOA** | 1.26 [1.13 - 1.41] | 5.14 × 10^-5^ | 1.31 [1.16 - 1.47] | 5.50 × 10^-6^ | 1.40 [1.20 - 1.65] | 3.21 × 10^-5^ | 1.29 [1.10 - 1.52] | 1.92 × 10^-3^ | 1.57 [1.34 - 1.85] | 3.17 × 10^-8^ | 1.32 [1.12 - 1.56] | 9.25 × 10^-4^ |
| **Odds of left knee rOA** | 1.33 [1.20 - 1.47] | 9.91 × 10^-8^ | 1.40 [1.25 - 1.56] | 1.28 × 10^-9^ | 1.41 [1.21 - 1.64] | 9.43 × 10^-6^ | 1.30 [1.12 - 1.52] | 6.58 × 10^-4^ | 1.75 [1.51 - 2.03] | 1.07 × 10^-13^ | 1.50 [1.29 - 1.74] | 1.84 × 10^-7^ |
| Right rKOA |  |  |  |  |  |  |  |  |  |  |  |  |
| **Odds of right hip rOA** | 1.31 [1.17 - 1.48] | 7.11 × 10^-6^ | 1.27 [1.13 - 1.44] | 1.28 × 10^-9^ | 1.29 [1.08 - 1.55] | 5.99 × 10^-3^ | 1.16 [0.97 - 1.40] | 0.11 | 1.65 [1.41 - 1.94] | 5.70 × 10^-10^ | 1.31 [1.12 - 1.55] | 1.07 × 10^-3^ |
| **Odds of left hip rOA** | 1.26 [1.13 - 1.41] | 5.14 × 10^-5^ | 1.32 [1.18 - 1.48] | 1.91 × 10^-6^ | 1.40 [1.20 - 1.65] | 3.21 × 10^-5^ | 1.29 [1.10 - 1.52] | 2.02 × 10^-3^ | 1.57 [1.34 - 1.85] | 3.17 × 10^-8^ | 1.31 [1.11 - 1.55] | 1.29 × 10^-3^ |
| **Odds of left knee rOA** | 32.80 [30.34 - 35.45] | <0.01 × 10^-324^ | 26.07 [24.07 - 28.24] | <0.01 × 10^-324^ | 39.32 [34.37 - 44.97] | <0.01 × 10^-324^ | 33.50 [29.21 - 38.43] | <0.01 × 10^-324^ | 27.81 [25.28 - 30.61] | <0.01 × 10^-324^ | 22.91 [20.77 - 25.27] | <0.01 × 10^-324^ |
| Left rKOA |  |  |  |  |  |  |  |  |  |  |  |  |
| **Odds of right hip rOA** | 1.41 [1.26 - 1.57] | 1.04 × 10^-9^ | 1.39 [1.24 - 1.55] | 1.32 × 10^-8^ | 1.38 [1.16 - 1.63] | 1.98 × 10^-4^ | 1.25 [1.06 - 1.49] | 9.16 × 10^-3^ | 1.79 [1.55 - 2.08] | 1.02 × 10^-14^ | 1.46 [1.25 - 1.70] | 1.17 × 10^-6^ |
| **Odds of left hip rOA** | 1.33 [1.20 - 1.47] | 9.91 × 10^-8^ | 1.41 [1.27 - 1.57] | 3.35 × 10^-10^ | 1.41 [1.21 - 1.64] | 9.43 × 10^-6^ | 1.30 [1.12 - 1.52] | 7.16 × 10^-4^ | 1.75 [1.51 - 2.03] | 1.07 × 10^-13^ | 1.49 [1.28 - 1.73] | 2.99 × 10^-7^ |
| **Odds of right knee rOA** | 32.80 [30.34 - 35.45] | <0.01 × 10^-324^ | 26.09 [24.09 - 28.26] | <0.01 × 10^-324^ | 39.32 [34.37 - 44.97] | <0.01 × 10^-324^ | 33.55 [29.25 - 38.49] | <0.01 × 10^-324^ | 27.81 [25.28 - 30.61] | <0.01 × 10^-324^ | 22.93 [20.79 - 25.30] | <0.01 × 10^-324^ |

*Adjusted for age, sex, race, height, weight and deprivation*

*rHOA – radiographic hip osteoarthritis, rKOA – radiographic knee osteoarthritis, OR – odds ratio, CI – confidence interval, P – p-value*

Supplementary Table 3 - Prevalence of radiographic osteoarthritis among the different quintiles of deprivation (TDI) and racial background

|  | **Right rHOA** | **Left rHOA** | **Right rKOA** | **Left rKOA** |
| --- | --- | --- | --- | --- |
|  | **Frequency [%]** | **Frequency [%]** | **Frequency [%]** | **Frequency [%]** |
| TDI quintiles |  |  |  |  |
| **1 (least deprived)** | 1803 [7.21] | 2,067 [8.27] | 1553 [6.21] | 1780 [7.12] |
| **2** | 969 [6.52] | 1,195 [8.04] | 948 [6.38] | 1068 [7.19] |
| **3** | 695 [6.88] | 765 [7.57] | 648 [6.41] | 686 [6.79] |
| **4** | 436 [6.62] | 559 [8.49] | 432 [6.56] | 485 [7.37] |
| **5 (most deprived)** | 195 [6.66] | 225 [8.71] | 169 [5.78] | 201 [6.87] |
| Race |  |  |  |  |
| **White** | 3,766 [6.87] | 4,430 [8.08] | 3,423 [6.24] | 3880 [7.08] |
| **Asian** | 43 [6.61] | 54 [8.29] | 44 [6.76] | 47 [7.22] |
| **Black** | 14 [3.54] | 35 [8.86] | 35 [8.86] | 40 [10.13] |
| **Mixed** | 11 [3.70] | 17 [5.72] | 22 [7.41] | 25 [8.42] |
| **Chinese** | 5 [3.25] | 5 [3.25] | 9 [5.84] | 12 [7.79] |
| **Unknown** | 259 [8.24] | 300 [9.55] | 217 [6.91] | 216 [6.87] |

*TDI – Townsend Deprivation Index, rHOA – radiographic hip osteoarthritis, rKOA – radiographic knee osteoarthritis*

Supplementary Table 4 - Binary logistic regression for the associations between radiographic osteoarthritis and quintiles of deprivation (TDI)

|  | **Unadjusted** | | **Fully Adjusted** | |
| --- | --- | --- | --- | --- |
|  | **OR [95% CI]** | **P** | **OR [95% CI]** | **P** |
| Right rHOA |  |  |  |  |
| **TDI quintile 2** | 0.90 [0.83 - 0.97] | 8.60 × 10^-3^ | 0.92 [0.85 - 1.00] | 0.05 |
| **TDI quintile 3** | 0.95 [0.87 - 1.04] | 0.27 | 1.01 [0.92 - 1.10] | 0.87 |
| **TDI quintile 4** | 0.91 [0.82 - 1.02] | 0.10 | 0.98 [0.88 - 1.10] | 0.75 |
| **TDI quintile 5** | 0.92 [0.79 - 1.07] | 0.28 | 1.02 [0.87 - 1.19] | 0.84 |
| Left rHOA |  |  |  |  |
| **TDI quintile 2** | 0.97 [0.90 - 1.04] | 0.42 | 0.99 [0.92 - 1.07] | 0.87 |
| **TDI quintile 3** | 0.91 [0.83 - 0.99] | 0.03 | 0.95 [0.87 - 1.04] | 0.26 |
| **TDI quintile 4** | 1.03 [0.93 - 1.13] | 0.56 | 1.09 [0.99 - 1.21] | 0.08 |
| **TDI quintile 5** | 1.06 [0.92 - 1.21] | 0.41 | 1.14 [0.99 - 1.31] | 0.07 |
| Right rKOA |  |  |  |  |
| **TDI quintile 2** | 1.03 [0.95 - 1.12] | 0.51 | 1.01 [0.93 - 1.10] | 0.74 |
| **TDI quintile 3** | 1.03 [0.94 - 1.14] | 0.48 | 1.03 [0.93 - 1.13] | 0.55 |
| **TDI quintile 4** | 1.06 [0.95 - 1.18] | 0.30 | 1.03 [0.92 - 1.16] | 0.56 |
| **TDI quintile 5** | 0.93 [0.79 - 1.09] | 0.35 | 0.91 [0.77 - 1.07] | 0.25 |
| Left rKOA |  |  |  |  |
| **TDI quintile 2** | 1.01 [0.93 - 1.09] | 0.81 | 0.99 [0.91 - 1.07] | 0.83 |
| **TDI quintile 3** | 0.95 [0.87 - 1.04] | 0.27 | 0.94 [0.85 - 1.03] | 0.18 |
| **TDI quintile 4** | 1.04 [0.93 - 1.15] | 0.49 | 1.01 [0.90 - 1.12] | 0.92 |
| **TDI quintile 5** | 0.96 [0.83 - 1.12] | 0.62 | 0.94 [0.80 - 1.09] | 0.41 |

*Adjusted for age, sex, race, height and weight*

*TDI – Townsend Deprivation Index, OR – odds ratio, CI – confidence interval, rHOA – radiographic hip osteoarthritis, rKOA – radiographic knee osteoarthritis, P – p-value*

Supplementary Table 5 – Binary logistic regression for the associations between radiographic osteoarthritis and racial groups

|  | **Unadjusted** | | **Fully Adjusted** | |
| --- | --- | --- | --- | --- |
|  | **OR [95% CI]** | **P** | **OR [95% CI]** | **P** |
| Right rHOA |  |  |  |  |
| **Asian** | 0.96 [0.70 - 1.31] | 0.79 | 1.19 [0.87 - 1.63] | 0.28 |
| **Black** | 0.50 [0.29 - 0.85] | 0.01 | 0.64 [0.37 - 1.10] | 0.11 |
| **Mixed** | 0.52 [0.29 - 0.95] | 0.03 | 0.71 [0.39 - 1.31] | 0.28 |
| **Chinese** | 0.46 [0.19 - 1.11] | 0.08 | 0.73 [0.30 - 1.79] | 0.49 |
| **Unknown** | 1.22 [1.07 - 1.39] | 3.00 × 10^-3^ | 1.15 [1.01 - 1.31] | 0.04 |
| Left rHOA |  |  |  |  |
| **Asian** | 1.03 [0.78 - 1.36] | 0.84 | 1.17 [0.88 - 1.56] | 0.28 |
| **Black** | 1.11 [0.78 - 1.57] | 0.57 | 1.32 [0.92 - 1.88] | 0.13 |
| **Mixed** | 0.69 [0.42 - 1.13] | 0.14 | 0.91 [0.56 - 1.50] | 0.72 |
| **Chinese** | 0.38 [0.16 - 0.93] | 0.03 | 0.58 [0.24 - 1.43] | 0.24 |
| **Unknown** | 1.20 [1.06 - 1.36] | 3.50 × 10^-3^ | 1.18 [1.04 - 1.34] | 0.01 |
| Right rKOA |  |  |  |  |
| **Asian** | 1.09 [0.80 - 1.48] | 0.59 | 1.65 [1.20 - 2.26] | 2.00 × 10^-3^ |
| **Black** | 1.46 [1.03 - 2.07] | 0.03 | 1.43 [0.99 - 2.06] | 0.05 |
| **Mixed** | 1.20 [0.78 - 1.86] | 0.41 | 1.29 [0.82 - 2.02] | 0.27 |
| **Chinese** | 0.93 [0.48 - 1.83] | 0.84 | 1.58 [0.80 - 3.14] | 0.19 |
| **Unknown** | 1.11 [0.97 - 1.28] | 0.14 | 0.90 [0.78 - 1.04] | 0.16 |
| Left rKOA |  |  |  |  |
| **Asian** | 1.02 [0.76 - 1.38] | 0.89 | 1.55 [1.14 - 2.10] | 5.20 × 10^-3^ |
| **Black** | 1.48 [1.07 - 2.06] | 0.02 | 1.43 [1.01 - 2.01] | 0.04 |
| **Mixed** | 1.21 [0.80 - 1.82] | 0.37 | 1.27 [0.83 - 1.95] | 0.26 |
| **Chinese** | 1.11 [0.62 - 2.00] | 0.73 | 1.89 [1.04 - 3.44] | 0.04 |
| **Unknown** | 0.97 [0.84 - 1.12] | 0.67 | 0.80 [0.69 - 0.92] | 2.30 × 10^-3^ |

*Adjusted for age, sex, height, weight and deprivation*

*White race as the comparator group*

*rHOA – radiographic hip osteoarthritis, rKOA – radiographic knee osteoarthritis, OR – odds ratio, CI – confidence interval, P – p-value*

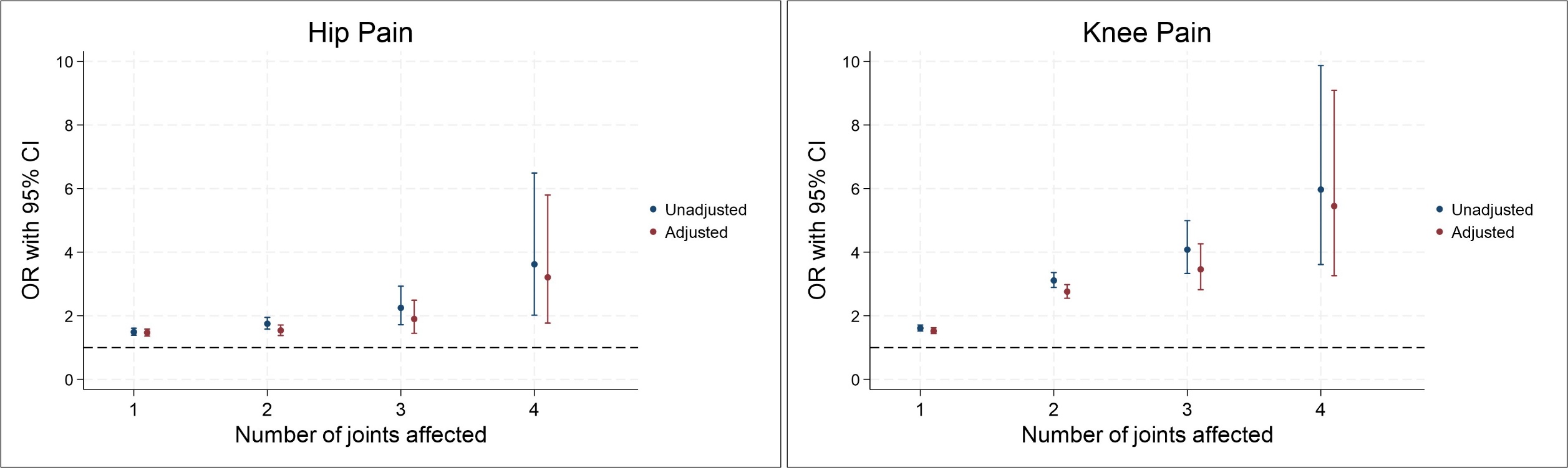

Supplementary Figure 1 - Logistic regression results for the associations between number of joints affected and self-reported hip or knee pain.

*Adjusted for age, sex, race, height, weight and deprivation*

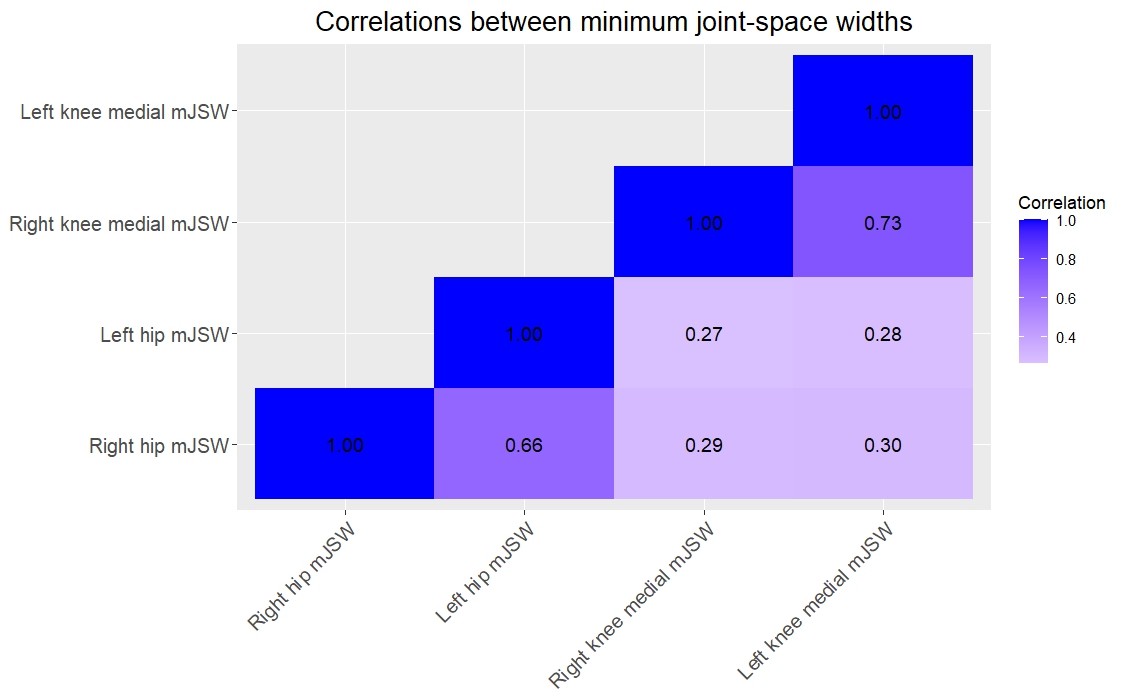

Supplementary Figure 2 - Correlations matrix to visually display the relationships between the minimum joint-space widths at each of the four anatomical joints

*mJSW- minimum joint-space width*
